# Hospital-Level Variation in Antenatal Corticosteroids for Late Preterm Births

**DOI:** 10.64898/2026.05.03.26351671

**Authors:** Mark A. Clapp, Dohyun Lee, Siguo Li, Kaitlyn E. James, Scott Lorch, Jessica L. Cohen, Jason D. Wright, Cynthia Gyamfi-Bannerman, Anjali J. Kaimal, Alexander Melamed

## Abstract

**Objective:** To determine whether and to what extent hospitals across the United States vary in their use of late-preterm steroids using a novel data set in which the timing of steroid administration relative to delivery can be observed.

**Methods:** This was a retrospective cohort study of singleton births with known gestational ages identified in the Premier Healthcare Database from 2015 to 2022. The primary variable of interest was hospital-level adoption of antenatal corticosteroids for late-preterm singleton deliveries, calculated as the proportion of late-preterm singleton births (34-36 completed weeks of gestation) with any betamethasone exposure during the same late-preterm period. Hospital adoption was defined as the weighted average rate of ALPS administration among late-preterm infants across the entire post-period. Hospitals were ranked by their late-preterm steroid adoption rates and categorized by quartile based on the empirical distribution. Temporal trends were assessed using annual hospital-level adoption rates and visualized using time-series plots and distributional plots. A logistic regression model was constructed to determine hospital characteristics associated with being a highest-quartile adopting hospital.

**Results:** The analysis cohort included 728 hospitals and 5,452,791 births, of which 361,006 (6.6%) were singleton late preterm births. Hospital steroid exposure rates ranged from 0 to 82% and were categorized into quartiles based on overall exposure rate, with cutoffs at 20.6%, 29.8%, and 40.1%. Median exposure rates increased progressively across quartiles from 14.1% (IQR 9.3-17.4%) in the lowest adopting hospitals (Q1) to 47.6% (IQR 43.7-53.2%) in the highest adopting hospitals (Q4), with substantial within-quartile variation. In the multivariable model, urban location was a strong predictor of high adoption after adjustment (aOR 2.05; 95% CI 1.11-3.83, p=0.02). Compared to Midwest hospitals, Southern hospitals had significantly lower odds of being high adopters (aOR 0.37; 95% CI 0.20-0.69, p<0.01). Among clinical case mix variables, a higher proportion of late preterm births at 34 weeks’ gestation was strongly associated with high adoption (aOR 2.21; 95% CI 1.58-3.14, p<0.001).

**Conclusion:** Following publication of the ALPS Trial, there was heterogeneous adoption of late preterm steroids among US hospitals. These findings highlight the need for a more in-depth exploration of local factors that drive the adoption of evidence-based practices outside of observable hospital characteristics.

## Introduction

In 2016, the Antenatal Late Preterm Steroid (ALPS) Trial was published and showed a 20% reduction in respiratory morbidity for neonates who were at risk for late preterm birth.^1^ This study expanded the existing evidence on the benefits of antenatal steroids for preventing respiratory complications among preterm births.^2^ After the trial was published, the American College of Obstetricians and Gynecologists (ACOG) and Society for Maternal-Fetal Medicine (SMFM) quickly adapted their clinical guidance and recommended antenatal steroids for those at risk for late preterm birth in populations similar to those included in the ALPS trial.^3,4^

Analyses using US birth certificate data demonstrated a rapid increase in antenatal steroid exposure among neonates born in the late preterm period.^5^ However, a follow-up analysis of the same data set revealed significant regional variation in the adoption of late preterm steroids that was not explained by observable patient or regional factors.^6^ As clinical policies and guidelines are often set locally (i.e., at the group or hospital level) and the US birth certificate data does not specify the gestational age of steroid exposure, our primary objective was to determine whether and to what extent hospitals across the United States vary in their use of late-preterm steroids using data set in which the timing of steroid administration relative to delivery can be observed. Second, we sought to determine whether hospital-level factors are associated with the observed variation, as this information may inform policy on benchmarking and discussions of appropriate and optimal use of antenatal steroids. Our hypothesis was that there would be significant hospital-level variation in adoption that would not be associated with observable hospital characteristics.

## Methods

This was a retrospective cohort study of singleton births with known gestational ages identified in the Premier Healthcare Database (PHD) from 2015 to 2022. The PHD includes nationally representative annual data for patient demographics, diagnoses, procedures, itemized billing records, medication charges (i.e., antenatal steroid use), and hospital characteristics.^7^

The ALPS trial was published online in February 2016 and in print in April 2016.^1^ Its findings informed major guideline updates with the Society for Maternal-Fetal Medicine (SMFM) and American College of Obstetricians and Gynecologists (ACOG) issuing revised clinical guidelines in August and October 2016, respectively.^1,3,4^ We assigned March 2016 to October 2016 to be the evidence dissemination and implementation period, August 2015 to October 2016 as the pre-period, and November 2016 to December 2022 as the post-period; these timelines were used in prior analyses.^5,6,8,9^

The primary variable of interest was hospital-level adoption of antenatal corticosteroids for late-preterm singleton deliveries, calculated as the proportion of late-preterm singleton births (34-36 completed weeks of gestation) with any betamethasone exposure during the same late-preterm period. Hospitals contributing at least the 15^th^ percentile of late-preterm deliveries (≥ 10.2 average annual late-preterm births) were included; this threshold was set a priori to reduce noise in the estimates from hospitals with very low late-preterm delivery volumes. Hospital adoption was defined as the weighted average rate of ALPS administration among late-preterm infants across the entire post-period. Hospitals were ranked by their late-preterm steroid adoption rates and categorized by quartile based on the empirical distribution. The variable definitions and classification methods are described in detail in the Supplement (eMethods).

Temporal trends were assessed using annual hospital-level adoption rates and visualized using time-series plots and distributional plots. To assess for movement across quartiles between the early post-period (2017-2018) and late post-period (2021-2022), mean adoption rates were calculated in the early and in the late post-period, ranked, and then categorized by quartile. A Sankey diagram was constructed to visualize the movement of hospitals between quartiles in the early and late post-period.

The following hospital-level variables were compared among the highest and lowest quartiles of adoption: geographic region (Midwest, Northeast, South, West; the most granular location variable), teaching status (teaching vs. non-teaching), urban vs. rural location, annual overall delivery volume, annual delivery volume by birth cohort (early preterm, late preterm, and term births), and the availability of Level III-IV neonatal care.^10^ Level III or IV neonatal care was assigned using a proxy based on the average annual number of inborn neonates delivered at <32 weeks’ gestation, identified using diagnosis codes for extreme prematurity or week-specific gestational age <32 weeks and linked to maternal delivery hospitalizations at the same hospital when infant admission dates occurred within 1 day of the maternal delivery date. Hospitals averaging 10 or more births at ≤32 weeks of gestation per year were classified as having Level III-IV NICU capability. More details on the neonatal designation of care are provided in the eMethods in the Supplement.

To account for potential differences in patient case mix, we calculated and compared hospital-level proportions of the following patient characteristics between the highest and lowest quartiles of adoption: percentage of preterm births occurring at 34 weeks’ gestation (vs. 35-36 weeks), percentage of late preterm births with a diagnosis of preterm labor or preterm premature rupture of membranes (i.e., spontaneous preterm births), percentage of late preterm births to multiple gestations, percentage of late preterm births with pre-existing type I or type II diabetes (individuals excluded from the ALPS Trial), and percentage of late preterm births with a diagnosis code for a history of preterm delivery.

Comparisons of hospital-level steroid exposure rates between groups were assessed using Wilcoxon rank-sum tests, given their skewed continuous distributions. Comparisons between the lowest and highest quartiles used analysis of variance for continuous variables and chi-squared tests for categorical variables. Also, a logistic regression model was constructed with the outcome of being a highest-quartile adopting hospital and the hospital characteristics outlined above as covariates.

We also conducted several sensitivity analyses. First, we used the same multivariable regression model to identify the characteristics of the highest adopters of antenatal steroids, excluding mothers who transferred facilities; this increased confidence that the observed steroid adoption rates were attributable to the clinical practices of the delivering facility rather than the protocols of the referral site. Second, as the lowest volume hospitals were excluded in the primary analysis (those with <10.2 late preterm births per year), we compared the hospital characteristics between those included and excluded in the analysis, and performed the same multivariable logistic regression model comparing characteristics between the highest and lowest quartile of adoption when those low volume hospital were included in the assignment of quartiles.

All analyses were performed in R version 4.4.2. P-values <0.05 were considered statistically significant. The Mass General Brigham IRB classified this as non-human subjects research as it used deidentified data.

## Results

The analysis cohort included 728 hospitals and 5,452,791 births, of which 361,006 (6.6%) were singleton late preterm births. Figure 1 shows the distribution of hospital-level rates of late preterm steroid exposure among late preterm births in the post-period (range 0 to 82%). Hospitals were categorized into quartiles based on their overall post-ALPS steroid exposure rate, with quartile cutoffs at 20.6%, 29.8%, and 40.1%. Median exposure rates increased progressively across quartiles from 14.1% (IQR 9.3-17.4%) in the lowest adopting hospitals (Q1) to 47.6% (IQR 43.7-53.2%) in the highest adopting hospitals (Q4), with substantial within-quartile variation.

**Figure 1:**
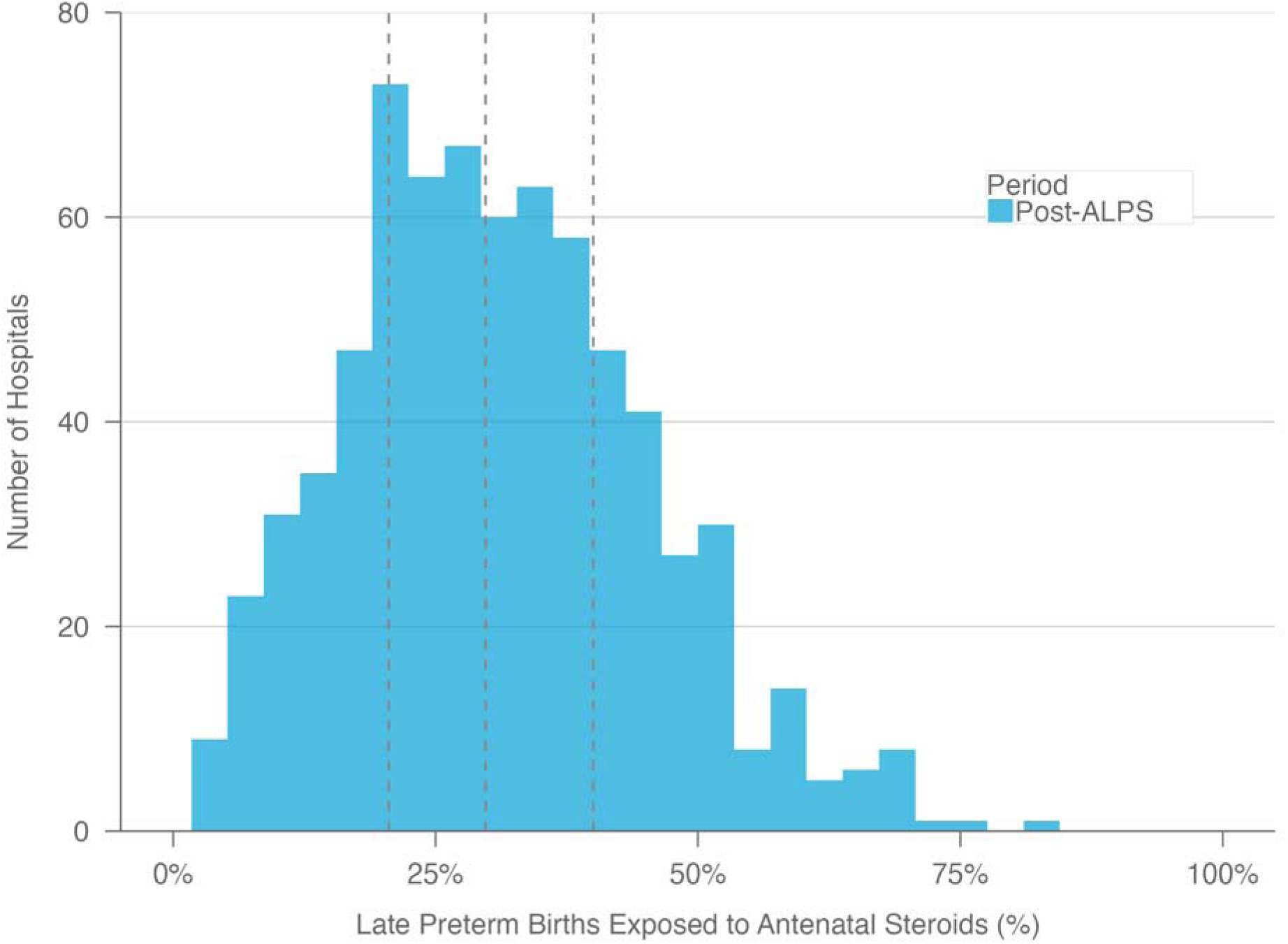
Distribution of Hospital Steroid Exposure Rates Among Late Preterm Births.

| Characteristic | Q1<br>(N = 182) | Q2<br>(N = 182) | Q3<br>(N = 182) | Q4<br>(N = 182) |
| --- | --- | --- | --- | --- |
| Overall Exposed (%) | 12.6 | 25.4 | 34.7 | 48.8 |
| Median Exposure (%) (IQR) | 14.1 (9.3-17.4) | 25.0 (22.5-27.4) | 35.1 (32.4-37.3) | 47.6 (43.7-53.2) |
| Range (%) | 0-20.5 | 20.6-29.7 | 29.8-40.0 | 40.1-82.1 |

The rates of late preterm births exposed to antenatal steroids were plotted over time and stratified by quartiles of adoption. The median hospital-level steroid exposure rate increased from 0.4% in the pre-ALPS period to 33.3% in 2022 (p<0.001). Figure 2A shows the monthly trends stratified by quartile of adoption from 2016 to 2018, shown with more granularity to demonstrate the pace of change after the ALPS Trial publication; Figure 2B shows the yearly trends across the entire analysis period.^1^ Steroid exposure increased across all quartiles following the 2016 ALPS Trial publication. Q4 hospitals demonstrated a sharp increase in steroid usage from 2016 to 2017 and remained elevated throughout the study period. In contrast, Q1 hospitals showed slower initial uptake and plateaued at lower rates, with steroid exposure rates remaining markedly divergent by 2022 (Q1: 14.3% vs Q4: 49.0% of late preterm births exposed to antenatal steroids, p<0.001) (Figure 2B).

**Figure 2:**
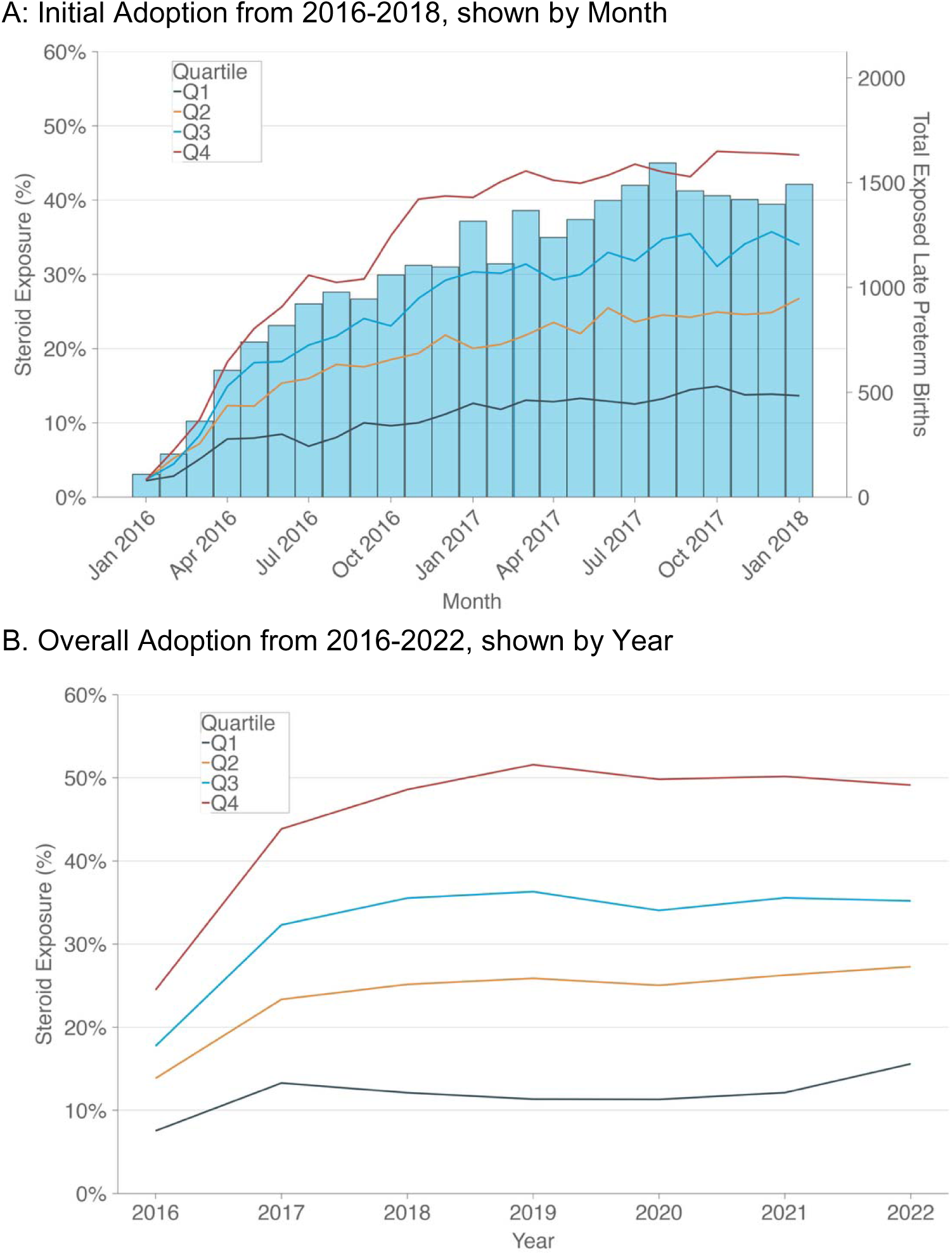
Trends in Steroid Exposure in Late Preterm Births by Quartile of Adoption. A) Monthly time series plot for steroid exposure among late preterm births in the first two years, stratified by quartile (Q). The right y-axis shows the total number of late preterm births exposed per month across all included hospitals. B) Yearly time series plot for steroid exposure among late preterm births over the analysis period, stratified by quartile (Q).

Hospitals varied significantly in characteristics across quartiles, as shown in Table 1. Q1 hospitals were more rural (Q1: 34.1% vs Q4: 12.1%, p<0.001), non-teaching (Q1: 68.7% vs Q4: 52.2%, p <0.001), and disproportionately located in the South (Q1: 51.1% vs Q4: 29.7%, p<0.001), compared to Q4 hospitals. Q4 hospitals had more births per month on average (Q1: 116.2 vs Q4: 145.3, p=0.04) and a higher proportion of late preterm births occurring at 34 weeks’ gestation (Q1: 12.1% vs Q4: 16.0%, p<0.001). Clinical characteristics were similar across quartiles, with no statistically significant differences in rates of PPROM, preterm labor, or history of preterm delivery. Q4 hospitals had a modestly higher proportion of patients with multiple gestations (Q1: 7.8% vs Q4: 8.5%, p=0.02) and Type I or II diabetes (Q1: 2.7% vs Q4: 3.2%, p<0.01), though the absolute differences were small.

**Table 1:**
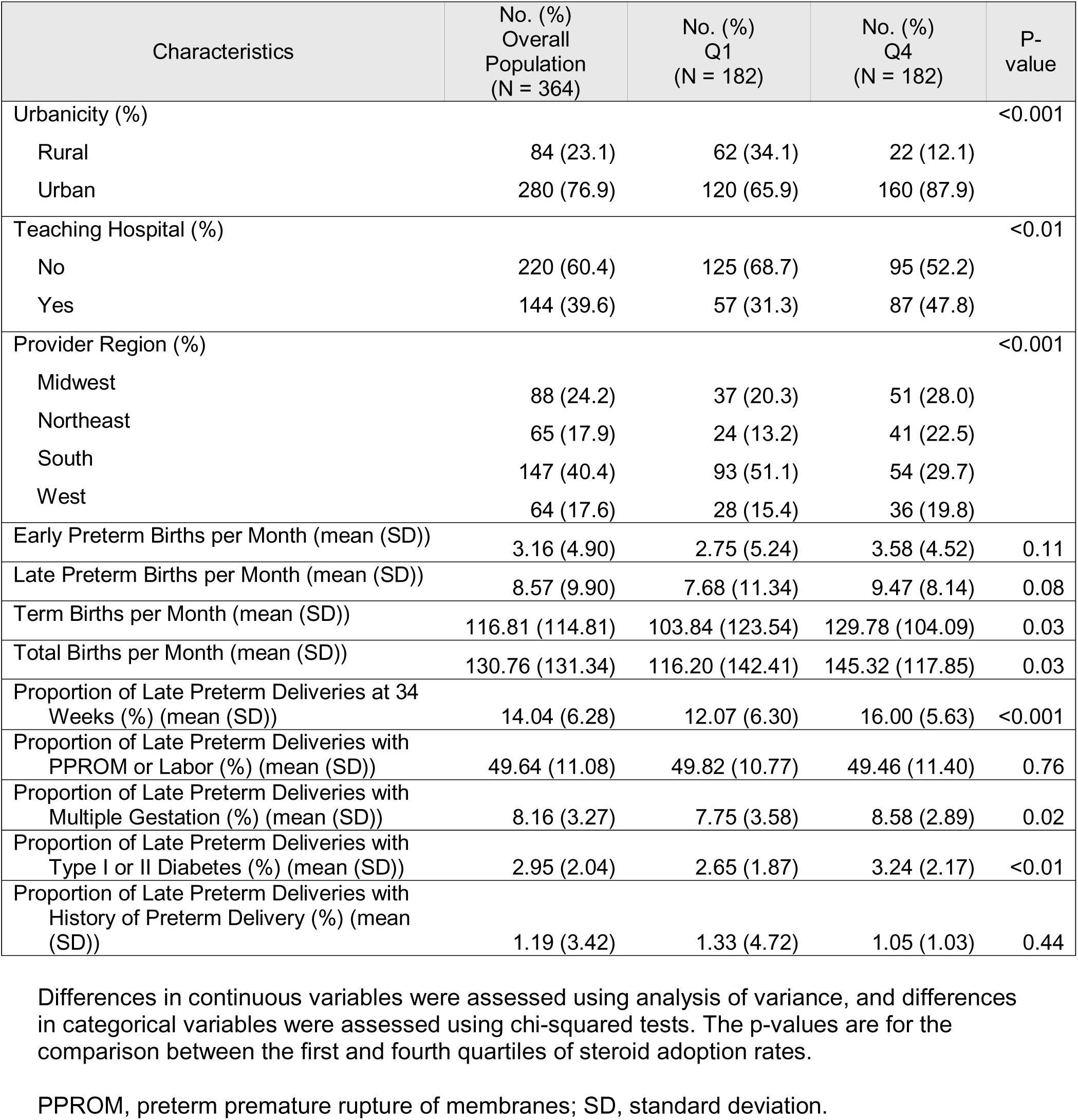
Characteristics of the Lowest and Highest Adopting Hospitals.

The Sankey diagram showing hospital quartile movement between the early and late post-periods is shown in Figure 3. Most of the lowest-adopting hospitals (Q1) in the early post-period remained low-adopting hospitals (Q1) in the post-period (n=59, 54.1%); only 7 (6.4%) moved from lowest to highest adopting hospitals across the time periods. Similarly, most of the highest-adopting hospitals (Q4) in the early post-period remained high adopters (Q4) over the time period (n=60, 55.6%), and 4 (3.7%) hospitals went from highest to lowest adopters between the early and late periods.

**Figure 3:**
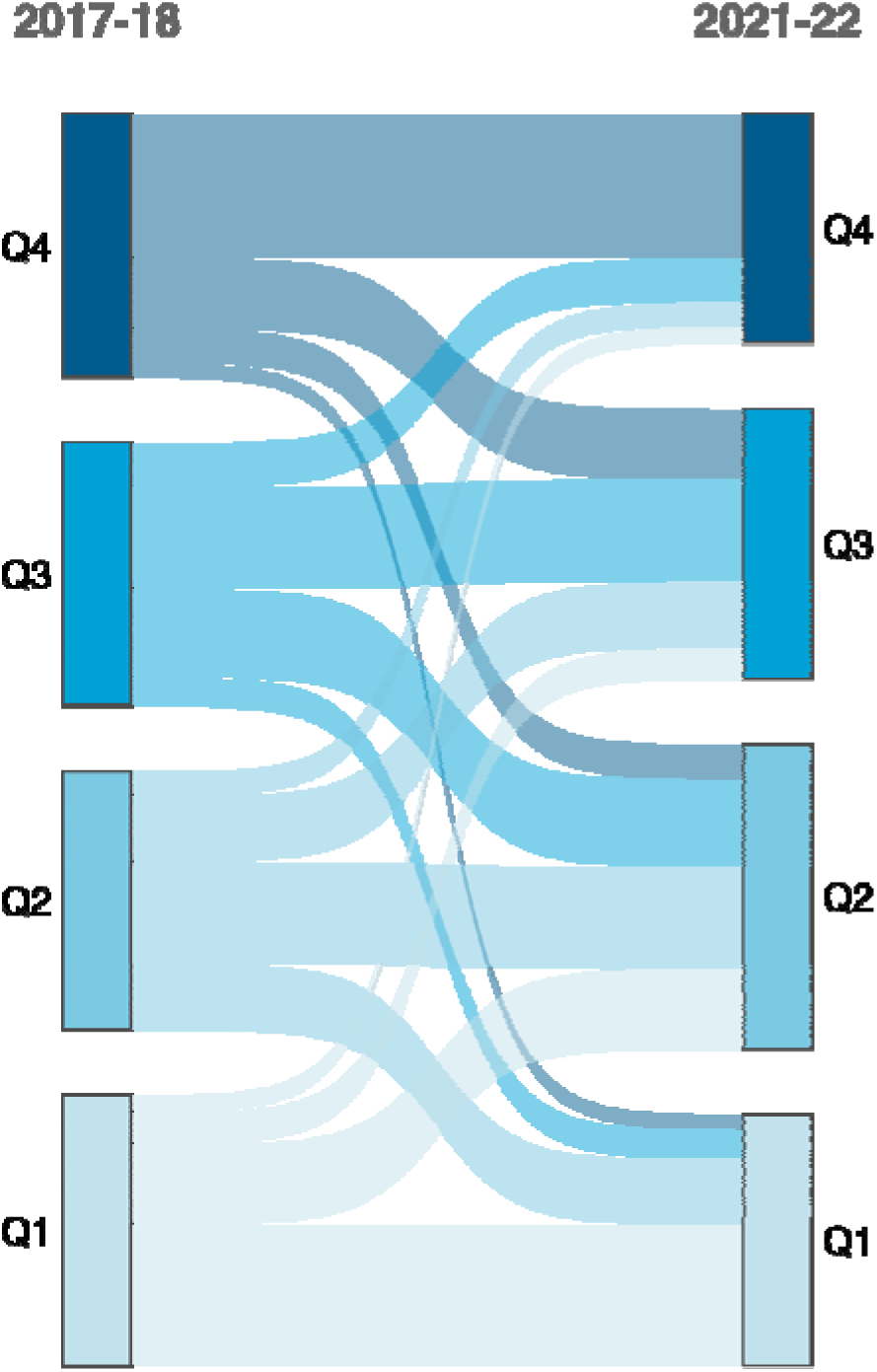
Sankey Diagram of Hospital Adoption Quartiles Between Early and Late Study Periods. Sankey diagram showing hospital movement across late-preterm steroid adoption quartiles between 2017-2018 and 2021-2022. Hospitals were assigned to quartiles based on their average adoption rate within each period, with thresholds calculated independently for each period. Flow width is proportional to the number of hospitals.

In the multivariable model, urban location was a significant predictor of high adoption after adjustment (aOR 2.05; 95% CI 1.11-3.83, p=0.02). Compared to Midwest hospitals, Southern hospitals had significantly lower odds of being high adopters (aOR 0.37; 95% CI 0.20-0.69, p<0.01). Among clinical case mix variables, a higher proportion of late preterm births at 34 weeks’ gestation was strongly associated with high adoption (aOR 2.21; 95% CI 1.58-3.14, p<0.001). Teaching status, birth volume, Level III or higher NICU designation, and higher proportions of PPROM or preterm labor, multiple gestation, and preterm delivery history were not independently associated with adoption quartile after adjustment. These findings are visually summarized in Figure 4. The unadjusted and adjusted odds ratios are shown in Supplemental Table 2.

**Figure 4:**
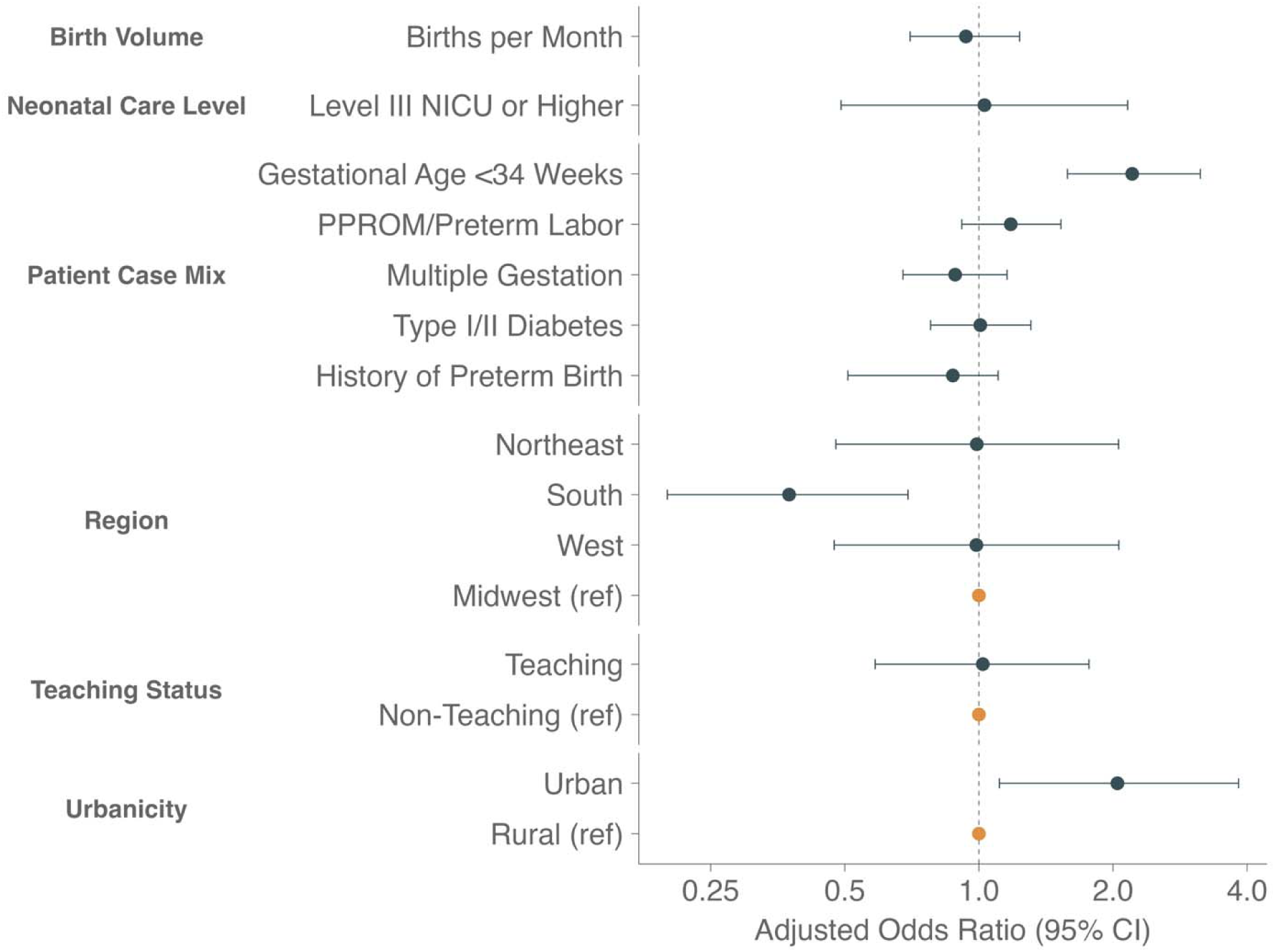
Hospital Factors Associated with High Adoption of Late Preterm Steroids. Adjusted odds ratios from logistic regression predicting the highest quartile adoption of antenatal corticosteroids for late preterm births.

The results from the sensitivity analysis excluding maternal transfers are shown in Supplemental Table 3. The findings were similar to those of the primary analysis. The characteristics between the excluded low-volume hospitals and the cohort included in the analysis are shown in Supplemental Table 4. The trend plots and results of the multivariable model when quartiles were assigned using all hospitals are shown in Supplement Figure 2 and Supplement Table 5. Marked variation between the highest- and lowest-quartile hospitals persisted, and hospital factors associated with the highest quartile remained similar.

## Discussion

Following the publication of the ALPS Trial, there was heterogeneous adoption of the practice of late preterm steroids among US hospitals, ranging from 0 to 82% of late preterm births being exposed to antenatal steroids. Trends in yearly exposure rates by adoption quartiles suggest that hospitals predominantly changed their practices within the first 1-2 years after the ALPS Trial, after which exposure rates in the quartiles remained constant through the end of the analysis period. The rate of steroid exposure among late preterm births in the highest quartile of adopting hospitals was 49% compared to approximately 13% in the lowest quartile.

Some hospital characteristics were associated with adoption: urban hospitals and hospitals with a higher percentage of late preterm births at 34 weeks of gestation were 2 times more likely to be high adopters, whereas hospitals in the South were about 3 times less likely to be high adopters (compared to Midwest hospitals). Other hospital and case-mix factors had weaker associations with adoption, suggesting that some degree of adoption was stochastic and not associated with measurable factors.

These findings highlight how clinical practice change in response to new evidence varies significantly, even when that new evidence is incorporated into clinical guidance issued by national professional societies, such as the ACOG and SMFM.^4,11^ Prior work has demonstrated significant regional variation (using the Dartmouth Atlas’s Healthcare Referral Regions) in steroid exposure among late preterm births (i.e., steroid exposure rates ranging from 0-47%), as reported on the US Birth Certificate, which has known issues with ascertainment for some data elements.^6,12,13^ In contrast, this hospital-level analysis is more granular, examining adoption at a unit of analysis that is more likely to reflect local practice patterns (i.e., hospital-based policies and/or culture) and was restricted to late preterm steroid exposure among late preterm births (compared to any prior steroid exposure among late preterm births). These findings highlight the need for a more in-depth exploration of local factors (e.g., local interpretation of evidence, culture, learning, champions, leadership) using structured frameworks (e.g., the Consolidated Framework for Implementation Research) that drive the adoption of evidence-based practices beyond observable hospital characteristics.^14–16^

In 2022, SMFM issued a statement proposing the use of antenatal steroid exposure as a hospital-based quality metric for obstetric care.^17^ This quality metric was focused on optimal antenatal steroid exposure among early preterm births (<34 weeks) and provided ideal and realistic hospital-level benchmarks. Late preterm births were excluded from that guidance, and the “ideal” or “optimal” rates of late preterm steroid administration are unknown, particularly as SMFM has issued guidance recommending shared decision-making around the practice.^17,18^

The data in this analysis may serve as a useful comparison for hospitals seeking to understand how their late-preterm steroid exposure rates compare with those of peer institutions in the absence of known or recommended benchmarks; for example, the histogram in Figure 1 can assist hospitals in identifying if their local practice is an outlier (either in low and high use of ALPS).

This study has several limitations. First, the Premier Healthcare Database represents a subset of U.S. hospitals and may not be fully generalizable to non-participating hospitals. Second, steroid identification relies on billing charge capture and may miss uncharged administrations or those occurring in other facilities prior to delivery hospitalization. The sensitivity analysis, which excluded maternal transfers and yielded the same results, reduced the risk of misattributing steroid administration. Third, gestational age is derived from ICD-10-CM Z3A codes, which provide week-level, but not day-level, resolution. To operationalize exposure windows, we assigned gestational age using a standardized midpoint-of-week assumption (e.g., 40 weeks coded as 40 weeks + 3 days). Although we use this approach to reduce classification bias, it may still lead to misclassification in the timing of steroid administration relative to delivery, particularly among patients near the bounds of the exposure window. Last, this was a hospital-level analysis that did not account for patient-level clinical factors that may influence the decision to administer steroids.

Following the ALPS Trial, there was heterogeneous adoption of late preterm steroids among US hospitals. After the initial adoption period, hospital-level rates of late preterm steroid exposure across quartiles remained nearly constant for >5 years, suggesting that local practice patterns became ingrained; these findings suggest to professional societies and policymakers that the opportunity to influence adoption is early and may become more challenging to effect change over time. Additional work is needed to understand local drivers of adoption and the degree to which ongoing clinical equipoise on the practice affects steroid use. Future studies comparing neonatal outcomes by adoption status may provide important data to catalyze practice change (either to increase or decrease utilization) and inform benchmarks and update policy and clinical guidance on the optimal use of late preterm steroids.

## Data Availability

Data sharing: not available.

## AI Disclosure

Claude (Anthropic, v1.1.7714) was used to identify areas of the manuscript where grammar, syntax, and/or readability could be improved after the manuscript was written. The authors take full responsibility for the accuracy and authenticity of the submitted work.

## References

1. Gyamfi-Bannerman C, Thom EA, Blackwell SC, et al. Antenatal Betamethasone for Women at Risk for Late Preterm Delivery. New England Journal of Medicine. 2016;374(14):1311–1320. doi:10.1056/NEJMoa1516783

2. Roberts D, Brown J, Medley N, Dalziel SR. Antenatal corticosteroids for accelerating fetal lung maturation for women at risk of preterm birth. Cochrane Database Syst Rev. 2017;3:CD004454. doi:10.1002/14651858.CD004454.pub3

3. American College of Obstetricians and Gynecologists’ Committee on Obstetric Practice, Society for Maternal– Fetal Medicine. Committee Opinion No.677: Antenatal Corticosteroid Therapy for Fetal Maturation. Obstet Gynecol. 2016;128(4):e187–194. doi:10.1097/AOG.0000000000001715

4. Society for Maternal-Fetal Medicine (SMFM) Publications Committee. Implementation of the use of antenatal corticosteroids in the late preterm birth period in women at risk for preterm delivery. Am J Obstet Gynecol. 2016;215(2):B13–15. doi:10.1016/j.ajog.2016.03.013

5. Clapp MA, Melamed A, Freret TS, James KE, Gyamfi-Bannerman C, Kaimal AJ. US Incidence of Late-Preterm Steroid Use and Associated Neonatal Respiratory Morbidity After Publication of the Antenatal Late Preterm Steroids Trial, 2015-2017. JAMA Netw Open. 2022;5(5):e2212702. doi:10.1001/jamanetworkopen.2022.12702

6. Freret TS, Cohen JL, Gyamfi-Bannerman C, et al. Regional Variation in Antenatal Late Preterm Steroid Use Following the ALPS Trial. JAMA Network Open. 2024;7(1):e2350830. doi:10.1001/jamanetworkopen.2023.50830

7. Data | Premier Products. Accessed April 3, 2020. https://products.premierinc.com/applied-sciences/solutions/data

8. Freret TS, James KE, Melamed A, Gyamfi-Bannerman C, Kaimal AJ, Clapp MA. Antenatal Steroid Exposure Among Term Newborns. JAMA Pediatr. 2022;176(12):1260–1261. doi:10.1001/jamapediatrics.2022.3251

9. Freret TS, James KE, Melamed A, Gyamfi-Bannerman C, Kaimal AJ, Clapp MA. Late-preterm steroid use among individuals with pregestational diabetes mellitus and with twin gestations. Am J Obstet Gynecol. Published online August 18, 2022:S0002-9378(22)00664-0. doi:10.1016/j.ajog.2022.08.029

10. American Academy of Pediatrics Committee on Fetus and Newborn. Levels of Neonatal Care. Pediatrics. 2012;130(3):587–597. doi:10.1542/peds.2012-1999

11. Practice Bulletin No. 171: Management of Preterm Labor □: Obstetrics & Gynecology. Accessed October 18, 2021. https://journals-lww-com.ezp-prod1.hul.harvard.edu/greenjournal/Fulltext/2016/10000/Practice_Bulletin_No__171__Management_of_Preterm.61.aspx

12. Landon BE, Keating NL, Barnett ML, et al. Variation in patient-sharing networks of physicians across the United States. JAMA. 2012;308(3):265–273. doi:10.1001/jama.2012.7615

13. Dietz P, Bombard J, Mulready-Ward C, et al. Validation of selected items on the 2003 U.S. standard certificate of live birth: New York City and Vermont. Public Health Rep. 2015;130(1):60–70. doi:10.1177/003335491513000108

14. Powell BJ, Mettert KD, Dorsey CN, et al. Measures of organizational culture, organizational climate, and implementation climate in behavioral health: A systematic review. Implement Res Pract. 2021;2:26334895211018862. doi:10.1177/26334895211018862

15. Fernandez ME, Walker TJ, Weiner BJ, et al. Developing measures to assess constructs from the Inner Setting domain of the Consolidated Framework for Implementation Research. Implement Sci. 2018;13(1):52. doi:10.1186/s13012-018-0736-7

16. Damschroder LJ, Aron DC, Keith RE, Kirsh SR, Alexander JA, Lowery JC. Fostering implementation of health services research findings into practice: a consolidated framework for advancing implementation science. Implement Sci. 2009;4:50. doi:10.1186/1748-5908-4-50

17. Hamm RF, Combs CA, Aghajanian P, Friedman AM. Society for Maternal-Fetal Medicine Special Statement: Quality metrics for optimal timing of antenatal corticosteroid administration. American Journal of Obstetrics and Gynecology. 2022;226(6):B2–B10. doi:10.1016/j.ajog.2022.02.021

18. Reddy UM, Deshmukh U, Dude A, Harper L, Osmundson SS. Society for Maternal-Fetal Medicine Consult Series #58: Use of antenatal corticosteroids for individuals at risk for late preterm delivery: Replaces SMFM Statement #4, Implementation of the use of antenatal corticosteroids in the late preterm birth period in women at risk for preterm delivery, August 2016. American Journal of Obstetrics & Gynecology. 2021;225(5):B36–B42. doi:10.1016/j.ajog.2021.07.023

